# Global, regional, and national burden of myocarditis and cardiomyopathy, 1990-2017

**DOI:** 10.1101/2020.09.08.20191007

**Authors:** Haijiang Dai, Dor Lotan, Arsalan Abu Much, Arwa Younis, Yao Lu, Nicola Luigi Bragazzi, Jianhong Wu

**Affiliations:** Centre for Disease Modelling, York University, Toronto, Ontario, Canada; Leviev Heart Center, Sheba Medical Center, Tel Hashomer; Sackler School of Medicine, Tel Aviv University, Tel-Aviv, Israel; Medical Screening Institute, Sheba Medical Center, Tel Hashomer, Tel-Aviv, Israel; Clinical Cardiovascular Research Center, University of Rochester Medical Center, Rochester, New York; Department of Cardiology, The Third Xiangya Hospital, Central South University, Changsha, China; Department of Health Sciences (DISSAL), Postgraduate School of Public Health, University of Genoa, Genoa, Italy

**Author notes:** **Corresponding authors:** Nicola Luigi Bragazzi, MD, PhD, MPH, Centre for Disease Modelling, York University, Address: 4700 Keele St, Toronto, Canada, M3J 1P3, (NLB), Yao Lu, MD, PhD, Department of Cardiology, The Third Xiangya Hospital, Central South University, Address: 138 Tongzipo Road, Changsha, China; 410013.

## Abstract

**Background:** To estimate the burden of myocarditis (MC) and cardiomyopathy for 195 countries and territories from 1990 to 2017.

**Methods:** We collected detailed information on MC and cardiomyopathy between 1990 and 2017 from the Global Burden of Disease study (GBD) 2017. Cardiomyopathy was divided into two types in GBD 2017, including alcoholic cardiomyopathy (AC) and other cardiomyopathy (OC). All estimates were presented as counts, age-standardised rates per 100 000 people and percentage change, with 95% uncertainty intervals (UIs).

**Results:** Worldwide, there were 1.80 million (95% UI 1.64 to 1.98) cases of MC, 1.62 million (95% UI 1.37 to 1.90) cases of AC and 4.21 million (95% UI 3.63 to 4.87) cases of OC, contributing to 46 486 (95% UI 39 709 to 51 824), 88 890 (95% UI 80 935 to 96 290) and 233 159 (95% UI 213 677 to 248 289) deaths in 2017, respectively. At the national level, the age-standardised prevalence rates varied by 10.4 times for MC, 252.6 times for AC and 38.1 times for OC; and the age-standardised death rates varied by 43.9 times for MC, 531.0 times for AC and 43.3 times for OC. Between 1990 and 2017, despite the decreases in age-standardised rates, the global numbers of prevalent cases and deaths have significantly increased for all the diseases. Females had greater decreases in age-standardised prevalence and death rates than males for all the diseases.

**Conclusions:** MC, AC and OC remain important global public health problems, and there are significant geographic variations in the burden for all these diseases. More effective and geo-specific strategies are necessary to counteract and mitigate the future burden of these diseases.

**Key questions:** *What is already known?:*

➢ Myocarditis (MC), alcoholic cardiomyopathy (AC) and other cardiomyopathy (OC) impose a substantial economic burden on healthcare systems. Studies that have systematically assessed the global, regional, and national burden of these diseases are still scarce.

*What are the new findings?:*

➢ Globally, there were an estimated 1.80 million (95% uncertainty interval (UI) 1.64 to 1.98) cases of MC, 1.62 million (95% UI 1.37 to 1.90) cases of AC and 4.21 million (95% UI 3.63 to 4.87) cases of OC in 2017.
➢ The global numbers of deaths due to MC, AC, and OC in 2017 were 46 486 (95% UI 39 709 to 51 824), 88 890 (95% UI 80 935 to 96 290) and 233 159 (95% UI 213 677 to 248 289), respectively.
➢ Across 21 world regions, the highest age-standardised prevalence rates of MC, AC and OC were seen in High-income Asia Pacific, Eastern Europe and Southern Sub-Saharan Africa, respectively. While the highest age-standardised death rates of MC, AC and OC were seen in Oceania, Eastern Europe and Central Europe, respectively.
➢ Despite the decreases in age-standardised rates, the global numbers of prevalent cases and deaths of MC, AC and OC have significantly increased between 1990 and 2017.

*What do the new findings imply?:*

➢ Our findings suggested that total numbers of prevalent cases and deaths of MC, AC and OC are increasing worldwide. More effective and geo-specific strategies aimed at counteracting and mitigating the future burden of these diseases are warranted.

## Introduction

Cardiovascular disease (CVD) is the largest single contributor to global mortality and morbidity, imposing a severe burden in terms of lost productivity and disability in adults.^1^ Among CVD, myocarditis (MC) represents an often-underdiagnosed cause of several life-threatening conditions, including acute heart failure, dilated cardiomyopathy and sudden death.^2 3^ The incidence and specific causes of MC vary considerably across the globe, depending on the geographical area and the selected population. In developed settings, MC is generally caused by viral pathogens, whereas in developing and resource-limited contexts, MC is mainly due to rheumatic carditis and infectious agents like *Trypanosoma cruzi* and diphtheria.^4^ Together with MC, cardiomyopathy is another often fatal disease contributing to about half of patients dying suddenly in childhood or adolescence or needing to undergo cardiac transplantation.^5^ Several types of cardiomyopathy exist, with new ones having been discovered and added recently and MC having been reclassified by some scholars as “inflammatory cardiomyopathy”.^5 6^

A thorough knowledge of the different epidemiological characteristics of MC and cardiomyopathy among various countries and populations can help public health decision- and policy-makers develop and implement targeted programs aimed at the prevention and management of the diseases. However, reliable and accurate epidemiological data regarding MC and cardiomyopathy are predominantly available from developed countries which apply well-established and consolidated diagnostic evaluations and criteria.^5^ The Global Burden of Disease study (GBD) is a continuous effort to evaluate the burden of more than 350 diseases and injuries for 195 countries and territories across the world and provides a unique opportunity to understand the landscape of MC and cardiomyopathy. Since 2016, cardiomyopathy was divided into two types in the GBD study, including alcoholic cardiomyopathy (AC) and other cardiomyopathy (OC).^7^ In the current study, we aimed to provide the estimates of prevalence, deaths, years lived with disability (YLDs), and years of life lost (YLLs) for MC, AC and OC for 195 countries and territories from 1990 to 2017, based on the most recent GBD 2017.

## Methods

### Overview

This study is part of GBD 2017, which is designed to provide a systematic assessment of health loss due to diseases and injuries at the global, regional, and national levels. In the latest iteration, GBD 2017, seven super-regions, 21 regions, and 195 countries and territories were included; and estimates for 359 diseases and injuries, 282 causes of death, and 84 risk factors were reported. The detailed methodology used for the estimation process can be found in previous GBD 2017 papers.^8-11^ Because GBD 2017 used de-identified, aggregated data, a waiver of informed consent was reviewed and approved by the University of Washington Institutional Review Board. All results and the original data sources used to produce estimates were publicly available on the website of Institute for Health Metrics and Evaluation, and can be accessed at http://ghdx.healthdata.org/.

### Case definitions

MC, AC and OC were ascertained using the International Classification of Diseases version 9 (ICD-9) and version 10 (ICD-10) codes. Diseases coded as 422-422.9 in ICD-9 or B33.2, I40-I41.9, and I51.4 in ICD-10 were identified as MC; diseases coded as 425.5 in ICD-9 or I42.6 in ICD-10 were identified as AC; and diseases coded as 425.0-425.3, 425.7-425.8, and 429.0 in ICD-9 or I42.1-I42.5, I42.7-I42.8, and I43-I43.9 in ICD-10 were identified as OC.^9^

### Estimation of deaths

We derived death estimates from vital registration data sources for MC and OC. We derived death estimates from vital registration and verbal autopsy data sources for AC. The ICD codes listed above were used to identify the diseases from the data sources. Data points that were implausibly high and discontinuous with the rest of the time series, or unable to be disaggregated appropriately were treated as outliers. Deaths due to MC, AC and OC were estimated using standard Cause of Death Ensemble model (CODEm), a highly systematised tool that runs many different models using location-level predictive covariates and chooses an ensemble of models that best reflects all the available input data.^10 12^ Predictive covariates incorporated into each cause-specific model can be found in the Table S1. To ensure consistency with all-cause mortality estimates, CoDCorrect algorithm was used to adjust the cause-specific results of MC, AC and OC for each age-sex-year-location group.

### Non-fatal estimation

The prevalence of MC, AC and OC was estimated using DisMod-MR 2.1, a Bayesian mixed-effects meta-regression tool developed for GBD analyses.^9^ Cause-specific incidence, remission, and mortality rates were integrated to produce consistent estimates of prevalence for all locations. Apart from the hospital and claims data, the prevalence estimates didn’t include any non-literature-based data. All outpatient data were excluded as they were implausibly low when compared with inpatient data from the same locations and claims data. For high-income North America, Central Europe, and Western Europe, any inpatient hospital data that were more than two-fold higher or 0.5-fold lower than the median absolute deviation value were also excluded for that age-sex group. More information on the prevalence estimation can be found elsewhere.^9^

YLDs were calculated by multiplying the prevalence (in number of cases) of each disease by the disability weight that quantifies the magnitude of heath loss associated with disease.^9^ Disability weight was measured on a scale from 0 to 1, where 0 represents full health and 1 is equivalent to death. Detailed explanation of the estimation process of disability weight has been described previously.^9^

### Socio-demographic Index

Socio-demographic Index (SDI) was a composite variable used to quantity the development level for each location-year.^8-11^ It ranged from 0 (less developed) to 1 (most developed), and was calculated based on lag distributed income per capita, mean education in the population aged 15 years or older, and total fertility rate under 25 years. 195 countries and territories in GBD 2017 were divided into five groups by SDI quintile: low SDI, low-middle SDI, middle SDI, high-middle SDI, and high SDI.^9^

### Compilation of results

YLLs were calculated by multiplying the number of deaths in each age group with a standard life expectancy at that age.^10^ The standard life expectancy was based on the lowest observed mortality risk for each age group in all populations over 5 million.^10^ Age-standardised rates were computed by direct standardization using the global age structure. Uncertainty was propagated throughout the modelling process by sampling 1000 draws at each step of the calculations. 95% uncertainty intervals (UIs) were determined as the 2.5th and 97.5th percentiles of the ordered 1000 draws.^8-11^ Any report on significant differences was based on a 95% UI excluding zero.

## Results

### Prevalence in 2017

Worldwide, there were 1.80 million (95% UI 1.64 to 1.98) cases of MC, 1.62 million (95% UI 1.37 to 1.90) cases of AC and 4.21 million (95% UI 3.63 to 4.87) cases of OC in 2017. The age-standardised prevalence rates of MC, AC and OC per 100 000 people in 2017 were 23.2 (95% UI 21.0 to 25.5), 19.9 (95% UI 16.8 to 23.3), and 53.9 (95% UI 46.7 to 62.1), respectively (Table 1). By sex, the age-standardised prevalence rate of AC was significantly higher in males than females [27.7 (95% UI 23.4 to 32.4) *vs*. 12.6 (95% UI 10.6 to 14.7) per 100 000 people], whereas the age-standardised prevalence rates of MC and OC were similar between males and females (Table 1). Figure 1 and Figure S1 showed the age-specific numbers and rates of prevalent cases by sex in 2017. In addition to extreme age groups, the numbers of MC prevalent cases were stable across different age groups in both sexes. The numbers of AC prevalent cases peaked at the ages of 65-69 years in both sexes, whereas the numbers of OC prevalent cases peaked at the ages of 65-69 years in males and 80-84 years in females.

**Figure 1.**
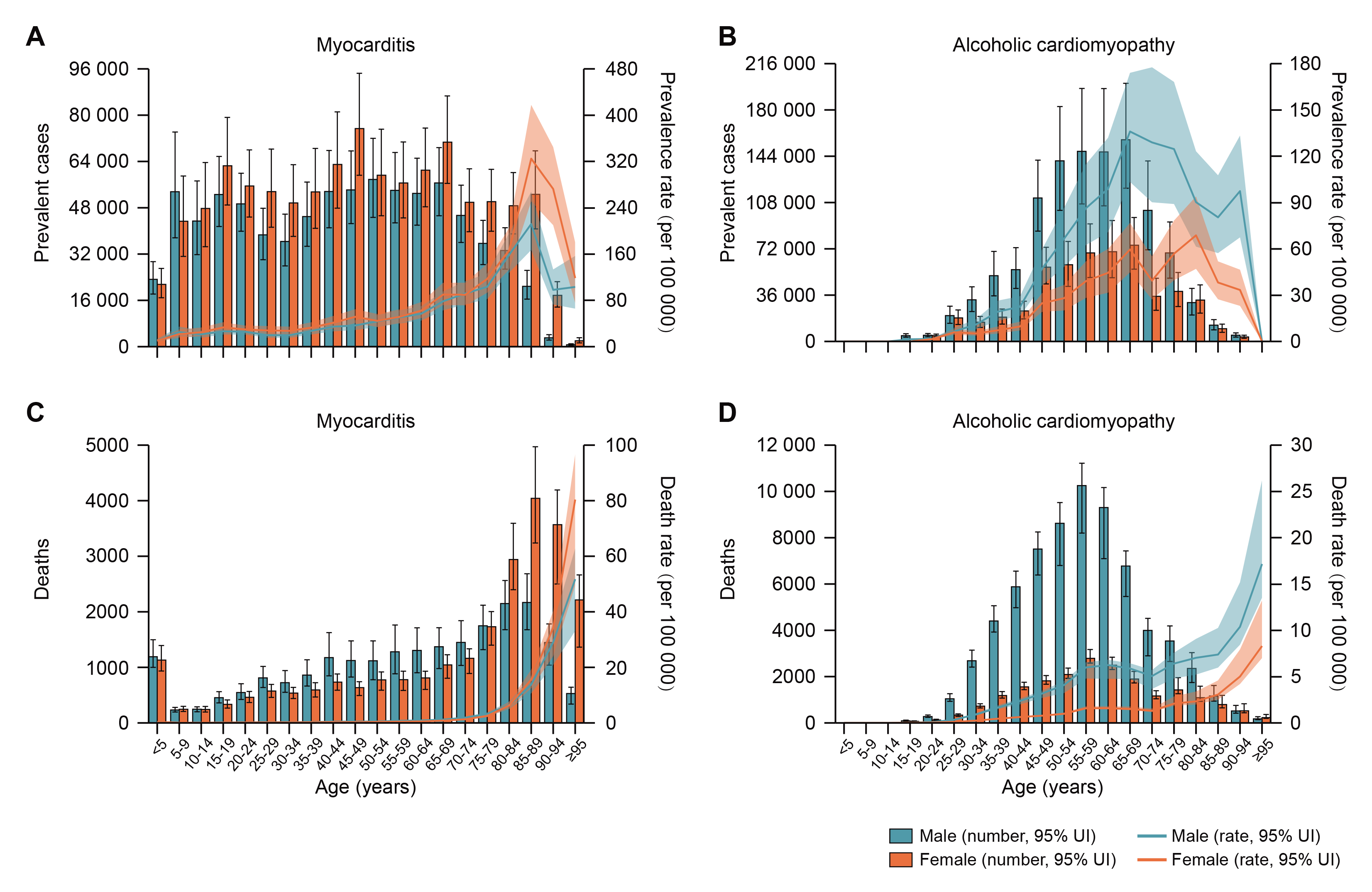
Age-specific numbers and rates of prevalent cases and deaths for myocarditis and alcoholic cardiomyopathy by sex, 2017. (A) Age-specific numbers and rates of myocarditis prevalent cases; (B) Age-specific numbers and rates of alcoholic cardiomyopathy prevalent cases; (C) Age-specific numbers and rates of myocarditis deaths; (D) Age-specific numbers and rates of alcoholic cardiomyopathy deaths.

**Table 1.** Age-standardised prevalence, death, YLD, and YLL rates of myocarditis, alcoholic cardiomyopathy, and other cardiomyopathy, and their percentage changes from 1990 to 2017, by sex and SDI quintile

|  | Prevalence |  | Deaths |  | YLDs |  | YLLs |  |
| --- | --- | --- | --- | --- | --- | --- | --- | --- |
|  | 2017<br>age-standardised<br>rate per 100 000<br>people | Percentage change in<br>age-standardised rates,<br>1990–2017 | 2017<br>age-standardised<br>rate per 100 000<br>people | Percentage change in<br>age-standardised rates,<br>1990–2017 | 2017<br>age-standardised<br>rate per 100 000<br>people | Percentage change in<br>age-standardised rates,<br>1990–2017 | 2017<br>age-standardised<br>rate per 100 000<br>people | Percentage change in<br>age-standardised rates,<br>1990–2017 |
| **Myocarditis** |  |  |  |  |  |  |  |  |
| Global | 23.2 (21.0 to 25.5) | -6.9% (-7.8 to -5.9) | 0.6 (0.5 to 0.7) | -6.2% (-15.0 to 3.4) | 1.7 (1.2 to 2.4) | -7.2% (-8.6 to -5.9) | 16.6 (14.5 to 18.5) | -23.5% (-32.6 to -11.4) |
| Sex |  |  |  |  |  |  |  |  |
| Male | 21.3 (19.4 to 23.4) | -4.7% (-5.6 to -3.7) | 0.7 (0.5 to 0.8) | 6.6% (-8.5 to 26.0) | 1.5 (1.0 to 2.1) | -4.7% (-6.1 to -3.2) | 18.3 (15.2 to 22.1) | -9.7% (-25.3 to 17.2) |
| Female | 24.9 (22.4 to 27.4) | -8.1% (-9.2 to -6.9) | 0.6 (0.5 to 0.6) | -15.6% (-24.4 to -0.3) | 1.9 (1.3 to 2.6) | -8.6% (-10.3 to -6.8) | 14.8 (12.7 to 16.7) | -35.5% (-44.9 to -22.8) |
| SDI quintile |  |  |  |  |  |  |  |  |
| Low SDI | 17.3 (15.8 to 19.0) | 1.6% (0.5 to 2.7) | 0.5 (0.3 to 0.6) | -16.6% (-30.7 to 2.7) | 1.2 (0.8 to 1.6) | 2.7% (0.2 to 5.1) | 14.6 (10.2 to 19.1) | -30.1% (-43.7 to -10.3) |
| Low-middle SDI | 19.6 (17.8 to 21.5) | 3.3% (2.2 to 4.3) | 0.6 (0.5 to 0.8) | -1.3% (-18.5 to 17.8) | 1.4 (0.9 to 1.9) | 4.3% (2.0 to 6.5) | 18.3 (14.7 to 22.5) | -22.0% (-36.6 to -2.6) |
| Middle SDI | 25.6 (23.0 to 28.3) | 3.1% (2.0 to 4.2) | 0.8 (0.6 to 0.9) | 17.6% (3.8 to 33.5) | 1.9 (1.3 to 2.6) | 4.4% (2.1 to 6.8) | 20.0 (16.8 to 23.5) | -19.8% (-30.6 to -5.0) |
| High-middle SDI | 25.7 (23.1 to 28.4) | 1.2% (-0.3 to 2.8) | 0.7 (0.6 to 0.8) | -14.2% (-26.9 to 7.0) | 1.9 (1.3 to 2.6) | 4.0% (0.9 to 7.4) | 17.0 (15.8 to 19.5) | -19.1% (-29.7 to -0.7) |
| High SDI | 27.1 (24.7 to 29.5) | -17.8% (-19.6 to -15.6) | 0.5 (0.4 to 0.5) | -13.1% (-36.9 to -1.5) | 2.1 (1.4 to 2.9) | -18.9% (-21.5 to -16.2) | 12.3 (10.1 to 13.2) | -14.8% (-35.6 to -5.1) |
| **Alcoholic**<br>**cardiomyopathy** |  |  |  |  |  |  |  |  |
| Global | 19.9 (16.8 to 23.3) | -26.2% (-28.2 to -24.0) | 1.1 (1.0 to 1.2) | -19.6% (-34.0 to -11.3) | 1.7 (1.2 to 2.4) | -25.7% (-28.0 to -23.2) | 34.7 (31.7 to 37.5) | -7.6% (-20.7 to 3.2) |
| Sex |  |  |  |  |  |  |  |  |
| Male | 27.7 (23.4 to 32.4) | -22.2% (-24.6 to -19.4) | 1.7 (1.5 to 1.9) | -6.2% (-18.7 to 6.6) | 2.4 (1.6 to 3.3) | -21.8% (-24.6 to -18.6) | 55.2 (47.0 to 60.4) | 3.3% (-14.4 to 17.4) |
| Female | 12.6 (10.6 to 14.7) | -33.6% (-35.4 to -31.6) | 0.5 (0.4 to 0.6) | -45.7% (-53.6 to -40.6) | 1.1 (0.7 to 1.5) | -32.8% (-35.2 to -30.1) | 14.8 (13.6 to 16.9) | -32.8% (-36.7 to -26.5) |
| SDI quintile |  |  |  |  |  |  |  |  |
| Low SDI | 11.8 (9.6 to 14.3) | 3.6% (1.0 to 6.2) | 0.4 (0.3 to 0.7) | -28.9% (-48.8 to -0.4) | 1.0 (0.7 to 1.5) | 4.2% (0.1 to 8.6) | 12.0 (7.8 to 17.8) | -31.5% (-49.8 to -5.1) |
| Low-middle SDI | 9.4 (7.8 to 11.3) | 5.8% (2.6 to 9.0) | 0.4 (0.3 to 0.5) | -22.3% (-41.3 to 1.9) | 0.8 (0.6 to 1.2) | 6.1% (0.4 to 12.1) | 11.5 (9.4 to 14.4) | -21.9% (-40.2 to 1.2) |
| Middle SDI | 7.1 (5.9 to 8.4) | 13.5% (11.5 to 15.5) | 0.3 (0.3 to 0.4) | 8.9% (-14.7 to 33.2) | 0.6 (0.4 to 0.9) | 13.5% (10.7 to 16.4) | 9.1 (7.4 to 11.1) | 12.3% (-11.5 to 34.0) |
| High-middle SDI | 27.2 (22.8 to 31.9) | -32.0% (-33.5 to -30.4) | 3.1 (2.9 to 3.5) | 5.9% (-14.5 to 31.2) | 2.3 (1.6 to 3.3) | -31.5% (-33.6 to -29.2) | 108.4 (101.1 to 120.6) | 19.6% (-3.8 to 52.9) |
| High SDI | 40.0 (34.5 to 46.1) | -17.7% (-21.7 to -12.9) | 0.8 (0.7 to 0.9) | -45.4% (-58.2 to -37.5) | 3.4 (2.3 to 4.7) | -17.7% (-22.5 to -12.5) | 22.8 (18.7 to 24.7) | -41.8% (-52.7 to -35.3) |
| **Other cardiomyopathy** |  |  |  |  |  |  |  |  |
| Global | 53.9 (46.7 to 62.1) | -15.5% (-18.4 to -12.7) | 3.1 (2.8 to 3.3) | -30.7% (-35.2 to -17.8) | 4.5 (3.1 to 6.3) | -15.0% (-17.8 to -12.1) | 71.1 (64.0 to 77.0) | -20.8% (-26.6 to -12.2) |
| Sex |  |  |  |  |  |  |  |  |
| Male | 53.1 (45.9 to 61.2) | -9.5% (-12.8 to -6.1) | 3.6 (3.1 to 3.9) | -22.9% (-30.5 to 7.8) | 4.5 (3.0 to 6.2) | -9.3% (-12.6 to -5.8) | 86.9 (77.0 to 95.1) | -11.9% (-21.0 to 3.5) |
| Female | 54.5 (47.1 to 62.6) | -18.7% (-21.6 to -15.9) | 2.6 (2.4 to 2.8) | -37.5% (-40.8 to -30.9) | 4.6 (3.1 to 6.3) | -18.1% (-21.1 to -15.1) | 55.7 (49.4 to 60.8) | -31.1% (-36.0 to -23.2) |
| SDI quintile |  |  |  |  |  |  |  |  |
| Low SDI | 48.6 (40.5 to 57.6) | 4.9% (2.2 to 7.5) | 3.0 (2.3 to 4.0) | -4.0% (-16.0 to 13.8) | 4.1 (2.7 to 5.8) | 5.6% (1.6 to 9.6) | 80.2 (61.5 to 105.9) | -15.5% (-28.0 to 0.6) |
| Low-middle SDI | 42.1 (35.5 to 49.5) | 7.6% (4.8 to 10.1) | 3.0 (2.6 to 3.4) | 4.0% (-10.6 to 23.0) | 3.5 (2.4 to 5.0) | 8.2% (4.4 to 11.9) | 76.3 (67.9 to 86.7) | -4.8% (-17.6 to 10.7) |
| Middle SDI | 32.2 (27.5 to 37.4) | 15.6% (13.8 to 17.6) | 2.1 (1.8 to 2.2) | 10.7% (0.9 to 25.5) | 2.7 (1.9 to 3.8) | 16.0% (13.7 to 18.5) | 50.6 (44.7 to 54.6) | -2.9% (-13.3 to 7.9) |
| High-middle SDI | 38.3 (33.0 to 44.3) | -3.6% (-6.2 to -0.8) | 3.7 (3.1 to 3.9) | 7.1% (-4.9 to 15.7) | 3.2 (2.2 to 4.5) | -2.5% (-5.7 to 0.7) | 88.0 (72.1 to 93.7) | 17.4% (-3.5 to 30.0) |
| High SDI | 97.8 (85.8 to 111.0) | -18.6% (-22.4 to -14.3) | 3.2 (3.0 to 3.4) | -51.0% (-55.0 to -33.2) | 8.2 (5.6 to 11.2) | -18.5% (-22.3 to -14.0) | 64.1 (61.2 to 74.7) | -45.2% (-49.5 to -26.4) |

Data in parentheses are 95% uncertainty intervals. YLDs= years lived with disability; YLLs= years of life lost; SDI= Socio-demographic Index.

The prevalence of MC, AC and OC in 2017 varied widely across geographic locations (Figure 2, Figure S2 and Table S2-S7). Regionally, High-income Asia Pacific had the highest age-standardised prevalence rate of MC [45.6 (95% UI 41.1 to 50.1) per 100 000 people], while the highest age-standardised prevalence rates of AC and OC were observed in Eastern Europe [115.6 (95% UI 97.1 to 135.8) per 100 000 people] and Southern Sub-Saharan Africa [150.3 (95% UI 120.5 to 184.0) per 100 000 people], respectively. Overall, national age-standardised prevalence rates of MC varied by 10.4 times across all countries, ranging from 10.2 (95% UI 9.0 to 11.4) per 100 000 people in Chile to 105.6 (95% UI 90.8 to 120.8) per 100 000 people in Albania (Figure 2 and Table S5). National age-standardised prevalence rates of AC varied by as much as 252.6 times, ranging from 1.4 (95% UI 1.2 to 1.7) per 100 000 people in Tajikistan to 356.1 (95% UI 300.9 to 419.6) per 100 000 people in Montenegro (Figure 2 and Table S6). Additionally, national age-standardised prevalence rates of OC varied by 38.1 times, ranging from 7.4 (95% UI 6.2 to 8.7) per 100 000 people in Kyrgyzstan to 280.3 (95% UI 236.2 to 330.6) per 100 000 people in Slovenia (Figure S2 and Table S7).

**Figure 2.**
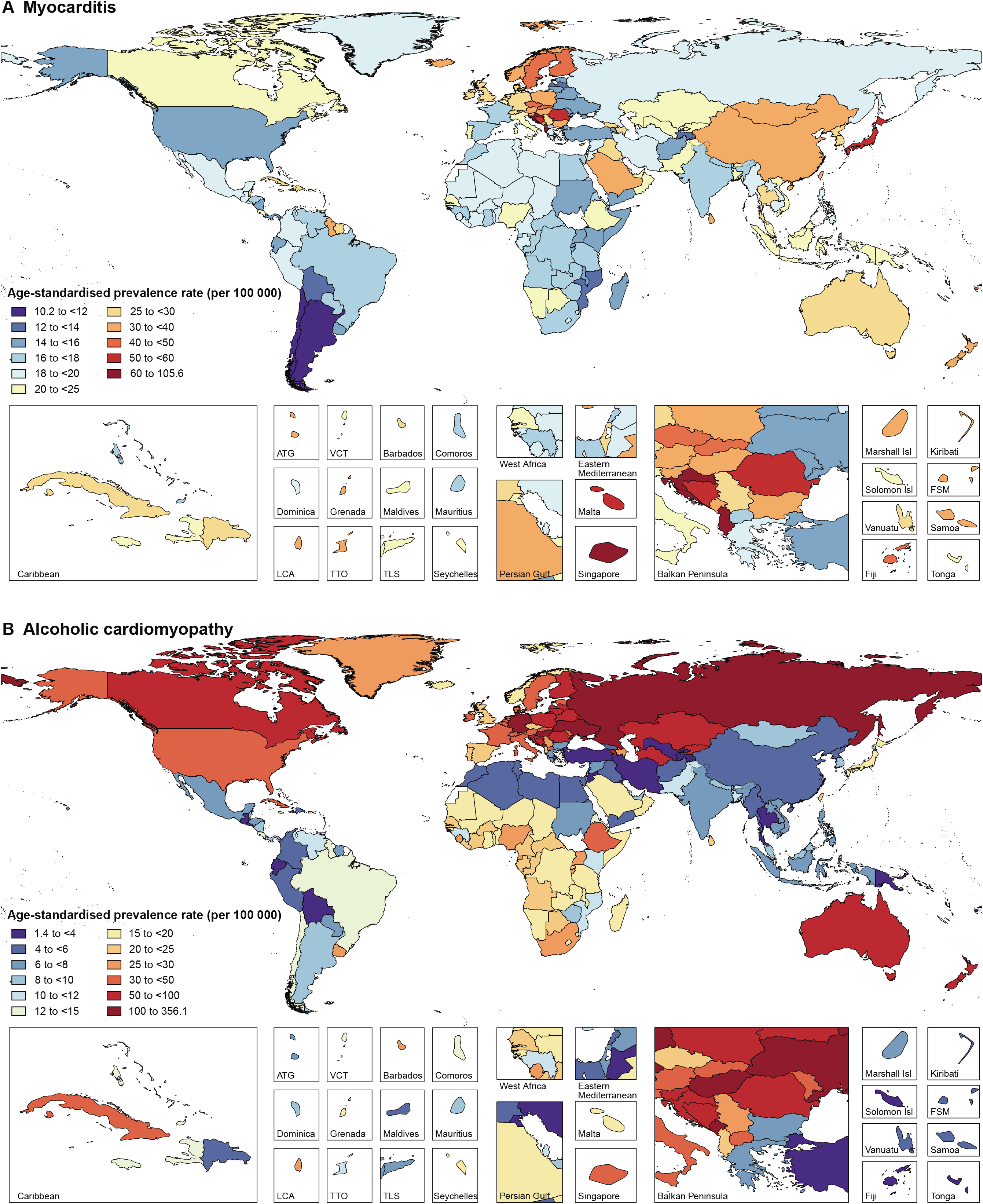
Age-standardised prevalence rates of myocarditis (A) and alcoholic cardiomyopathy (B) for 195 countries and territories, both sexes, 2017. ATG, Antigua and Barbuda; Isl, Islands; FSM, Federated States of Micronesia; LCA, Saint Lucia; TLS, Timor-Leste; TTO, Trinidad and Tobago; VCT, Saint Vincent and the Grenadines.

### Deaths, YLDs and YLLs in 2017

In 2017, the global numbers of deaths attributable to MC, AC and OC were 46 486 (95% UI 39 709 to 51 824), 88 890 (95% UI 80 935 to 96 290) and 233 159 (95% UI 213 677 to 248 289), respectively. The age-standardised death rates of MC, AC and OC per 100 000 people in 2017 were 0.6 (95% UI 0.5 to 0.7), 1.1 (95% UI 1.0 to 1.2), and 3.1 (95% UI 2.8 to 3.3), respectively (Table 1). By sex, the age-standardised death rate of MC was similar between males and females, whereas the age-standardised death rates of AC and OC were higher in males than females (Table 1). By age group, the numbers of deaths peaked at the ages of 85-89 years for MC and 55-59 years for AC in both sexes, while the numbers of deaths attributable to OC peaked at the ages of 80-84 years in males and 85-89 years in females (Figure 1 and Figure S1). Notably, the numbers of deaths attributable to MC and OC were much higher in children aged < 5 years than other young people.

Across 21 world regions, Oceania had the highest age-standardised death rates of MC [2.6 (95% UI 2.0 to 3.4) per 100 000 people], whereas the highest age-standardised death rates of AC and OC were seen in Eastern Europe [17.2 (95% UI 16.2 to 19.1) per 100 000 people] and Central Europe [10.4 (95% UI 9.6 to 11.0) per 100 000 people], respectively (Figure 3). At the national level, the age-standardised death rates of MC varied by 43.9 times across all countries, ranging from 0.10 (95% UI 0.08 to 0.12) per 100 000 people in Bahrain to 4.3 (95% UI 3.4 to 5.4) per 100 000 people in Albania (Figure S3 and Table S5). National age-standardised death rates of AC varied by 531.0 times, ranging from 0.04 (95% UI 0.03 to 0.04) per 100 000 people in Turkey to 18.7 (95% UI 17.3 to 21.5) per 100 000 people in Russian Federation (Figure S4 and Table S6). National age-standardised death rates of OC varied by 43.3 times, ranging from 0.41 (95% UI 0.33 to 0.47) per 100 000 people in Kyrgyzstan to 17.6 (95% UI 15.5 to 19.3) per 100 000 people in Dominica (Figure S5 and Table S7).

**Figure 3.**
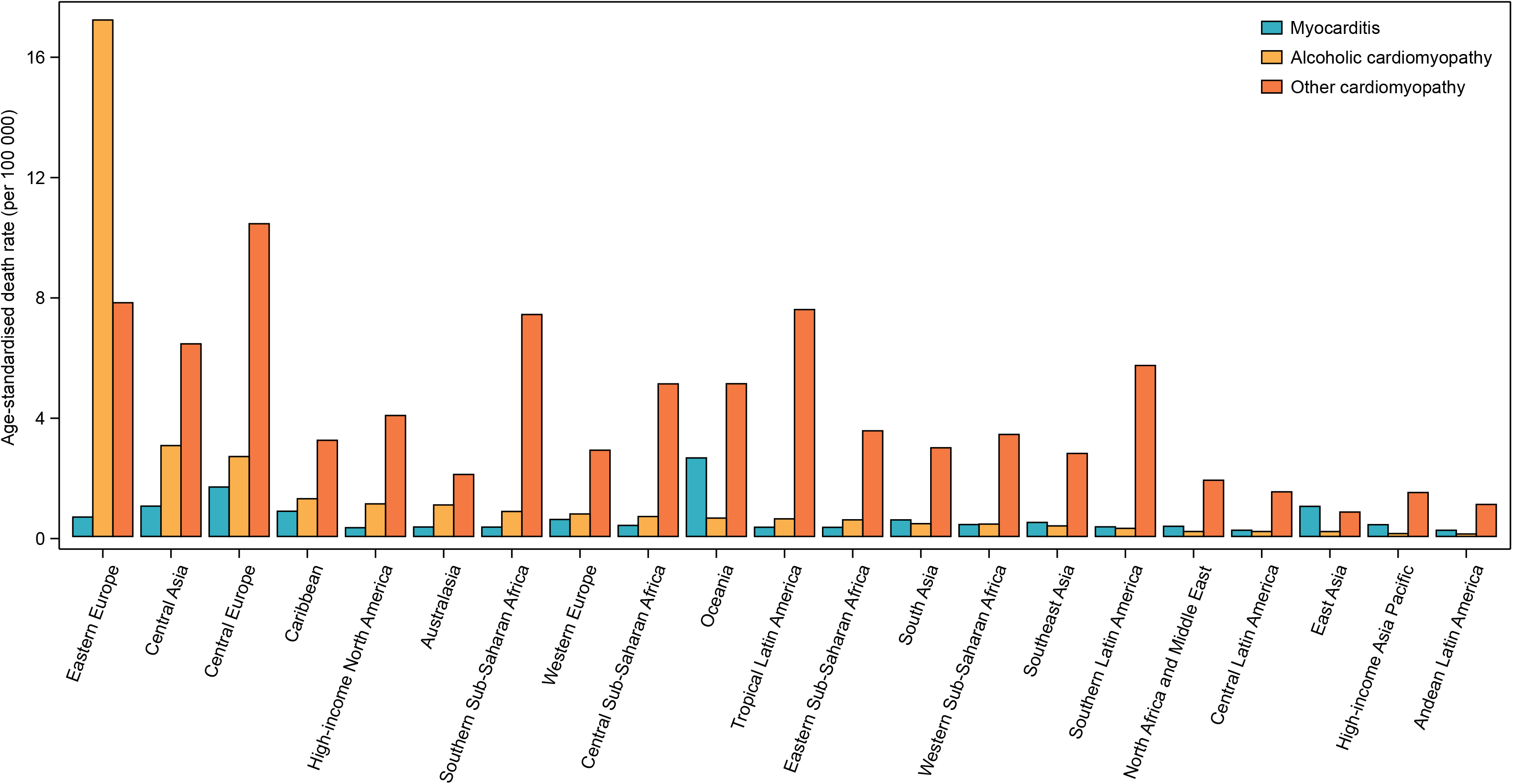
Age-standardised death rates of myocarditis, alcoholic cardiomyopathy, and other cardiomyopathy for 21 world regions, both sexes, 2017

Moreover, globally, there were 131 376 (95% UI 90 113 to 183 001) YLDs and 1.26 million (95% UI 1.10 to 1.42) YLLs attributable to MC, 139 087 (95% UI 95 134 to 196 130) YLDs and 2.84 million (95% UI 2.60 to 3.07) YLLs attributable to AC, and 353 325 (95% UI 237 907 to 493 908) YLDs and 5.51 million (95% UI 4.95 to 5.99) YLLs attributable to OC in 2017. The age-standardised YLD and YLL rates of these diseases by sex, SDI quintile, and geographic location can be seen in Table 1 and Table S5-S7.

### Temporal Trends of Burden from 1990 to 2017

Between 1990 and 2017, the age-standardised prevalence rates of MC, AC and OC decreased by −6.9% (95% UI −7.8 to −5.9), −26.2% (95% UI −28.2 to −24.0), and −15.5% (95% UI −18.4 to −12.7), respectively, with greater decreases in females than males for all the diseases (Table 1). Despite the decreases, the global numbers of MC, AC and OC have increased by 49.8% (95% UI 45.7 to 54.0), 39.0% (95% UI 35.3 to 43.1), and 65.3% (95% UI 60.2 to 70.8), respectively. By SDI quintile, the greatest decreases in age-standardised prevalence rates were found in countries in the high SDI quintile for both MC and OC, but in countries in the high-middle SDI quintile for AC (Table 1).

Similarly, the global numbers of deaths attributable to MC, AC and OC have increased by 71.4% (95% UI 54.9 to 90.5), 59.6% (95% UI 33.9 to 76.3), and 49.7% (95% UI 39.5 to 73.4), respectively from 1990 to 2017, despite the decreases in age-standardised death rates (Table 1). The decreases in age-standardised death rates were also greater in females than males for MC, AC and OC (Table 1). However, the change patterns in age-standardised death rates across SDI quintiles were not similar to age-standardised prevalence rates for all the diseases (Table 1). Furthermore, there were no clear patterns between age-standardised death rates for the 21 world regions and SDI over the 1990-2017 period for all the diseases (Figure 4 and Figure S6). These results indicated that the SDI level of the country is not a major determinant of size of deaths for these diseases. More information about the temporal changes in the burden of MC, AC and OC from 1990 to 2017 was available in Table 1 and Table S5-S7.

**Figure 4.**
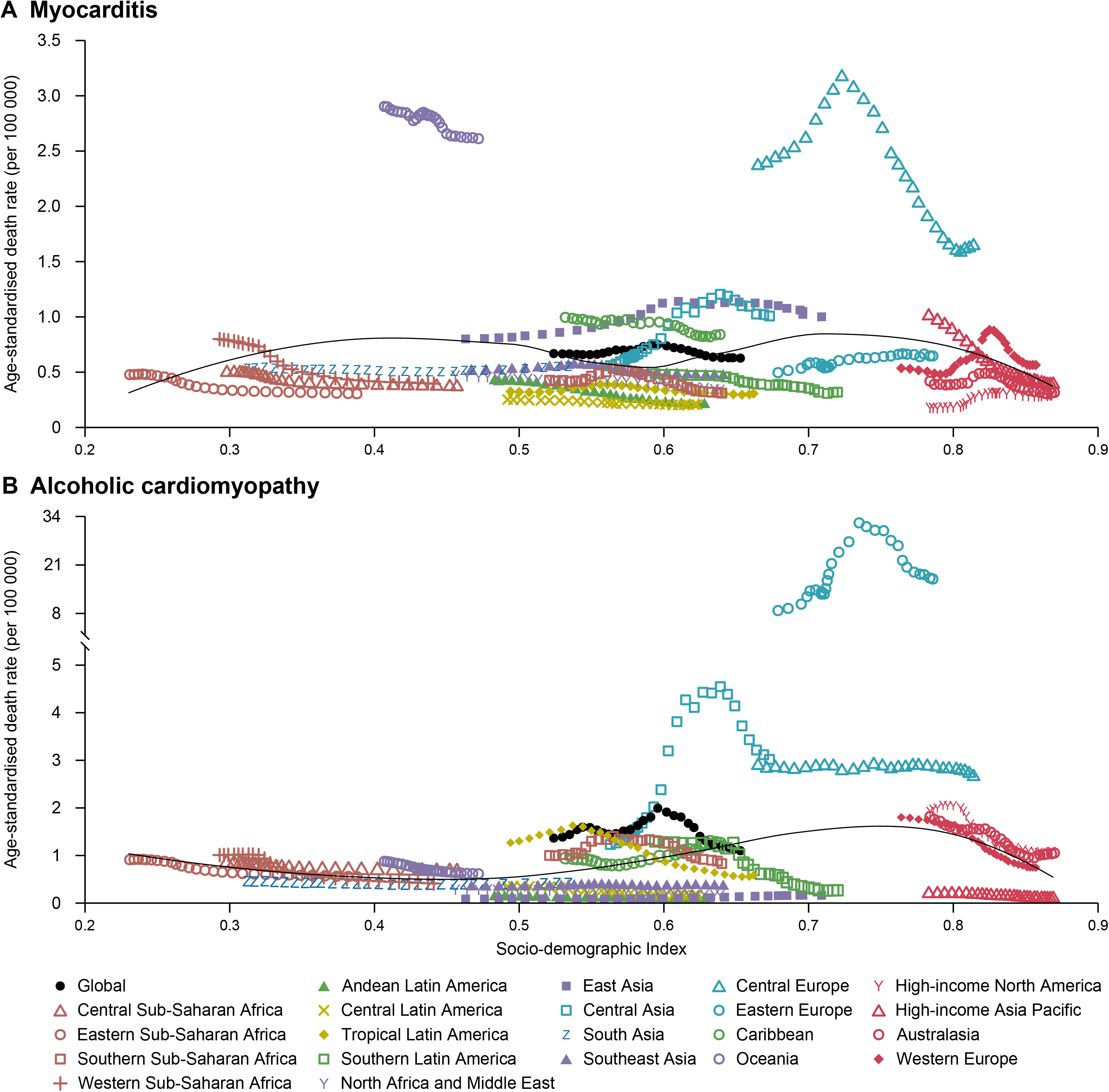
Temporal trends in age-standardised death rates of myocarditis (A) and alcoholic cardiomyopathy (B) for 21 world regions by SDI, both sexes, 1990-2017. For each region, points from left to right depict estimates from each year from 1990 to 2017. SDI = Socio-demographic Index.

## Discussion

In this study, we exploited the GBD 2017 modeling framework to estimate the global burden generated by MC, AC and OC, stratifying findings by age, sex, year (1990-2017), SDI, and geographic location. Most recently, a modelling study has estimated the death rate of AC in 2015 at the global, regional, and national levels, but without examining the prevalence, YLDs, and YLLs or exploring temporal trends of the burden over time.^13^ Additionally, to the best of our knowledge, no prior study has systematically reported the burden of MC and OC at the global, regional, and national levels. In the present study, we found that there were an estimated 1.80 million cases of MC, 1.62 million cases of AC and 4.21 million cases of OC globally in 2017, contributing to 46 486, 88 890 and 233 159 deaths, respectively. This study provides, indeed, health decision- and policy-makers with an updated synthesis of the epidemiology of MC, AC and OC to inform the strategic development, planning of preventative and treatment programs and optimization of health system resource allocation.

As expected, the age-standardised prevalence and death rates of AC were found to be significantly higher in males than females, and the burden of AC mainly concentered in adults aged 40-70 years. Alcohol consumption represents a major risk factor for global disease burden and generates substantial health losses.^14^ The higher frequency of alcohol use, as the risk factor for AC, in males compared with females can explain the higher prevalence rate of AC in males.^15^ Considering the significant impact of alcohol intake as etiopathogenetic driver of AC, the analysis of national trends and projections of alcohol consumption remain essential for AC surveillance and the development of targeted prevention strategies. In addition, it is noteworthy that although the age-standardised prevalence rates of MC and OC were lowest in children aged < 5 years, their corresponding numbers of deaths were much higher than in other young people. Actually, pediatric cardiomyopathies have a poor prognosis despite optimal medical management, with 40% of children with symptomatic cardiomyopathy dying or undergoing cardiac transplantation within 2 years.^16^ Further studies are warranted in order to fully understand the risk factors of pediatric cardiomyopathies to improve prognosis.

The burden of MC in 2017 varied significantly across geographic locations. Regionally, High-income Asia Pacific had the highest age-standardised prevalence rate of MC, which is probably because of the infection caused by hepatitis C virus (HCV).^17^ According to a study by Matsumori and colleagues,^18^ anti-HCV antibodies are frequently detected in MC patients with a higher rate than in the general population. While among all the industrialized countries, Japan has the highest prevalence rate of HCV, with more than 1 million cases of infections.^19^ However, despite High-income Asia Pacific reported the highest age-standardised prevalence rate of MC, the highest age-standardised death rate due to MC was seen in Oceania, which is probably due to the lack of adequate healthcare resources, especially in remote underserved areas.

In addition, large geographic differences in the burden of AC and OC were also observed in the present study. The highest age-standardised prevalence rates of AC in Eastern Europe may be explained taking into account the high level of alcohol consumption, in terms of drinking pattern, frequency, quality and type of alcoholic beverage. It has been reported that the detrimental effects of drinking patterns such as alcohol binge drinking are a major public health issue, in Europe and especially in Eastern Europe.^20^ Even though these countries have enacted restrictive policies on alcohol consumption which have contributed to a decline in alcohol-related mortality rate, the burden remains dramatically high.^21^ The highest age-standardised prevalence rate of OC in Southern Sub-Saharan Africa can be partly explained by the high prevalence of communicable disease, in particular infectious agents like *Toxoplasma gondii* and *Schistosoma*.^22^ Also, peripartum cardiomyopathy is quite widespread in the African continent.^22 23^ On the other hand, in resource-limited regions like Sub-Saharan Africa, the true prevalence of OC could even be under-estimated, due to the lack of detection systems.

Concerning the temporal trends of burden from 1990 to 2017, the global numbers of prevalent cases and deaths for MC, AC and OC have significant increased, contrasting with the decreases in age-standardized prevalence and death rates. The exact causes for the trends are unknown but may be in part due to population growth and aging.^24^ In terms of sex, females had greater decreases in age-standardised prevalence and death rates than males for all the diseases during 1990-2017, suggesting that preventive policies and intervention treatments may be more effective in females in the same timeframe. To our best knowledge, the associations of the burden of MC, AC and OC with developing status of regions and countries haven’t been explored in previous studies. In the present study, we found that there were no clear patterns between age-standardised death rates for the 21 world regions and SDI over the 1990-2017 period for all the diseases, suggesting that such associations are complex and nonlinear. Indeed, the burden of MC, AC and OC is not limited to developed or less developed countries and a high burden of these diseases was seen in countries with various SDI.

## Limitations

Even though the GBD estimations fill a gap where data on the burden imposed by MC, OC, and AC are scarce or inaccessible, several limitations still exist and should be properly acknowledged. The first limitation is the low availability of data in some countries, although statistically robust approaches have been applied in order to overcome data scarcity and deal with uncertainty. In those countries for which data was missing or was deemed inaccurate and of low quality, results mainly relied on covariates known to be associated with these diseases, trends in neighboring countries, or a combination of both methods. Besides, differences in data collection and data sources quality across countries, delay and inaccuracy in reporting, misclassification and bias in coding, are unavoidable in this study, even though the GBD has attempted to enhance the reliability and comparability of related data. Moreover, diagnostic criteria may have changed over time and may reflect di erent regional use of coding. Various nomenclatures, classification schemes and nosological taxonomies, indeed, exist, some of them being even confusing and contrasting, and have been differently employed in the literature.^25^

## Conclusions

In summary, MC, AC and OC remain important global public health problems; however, large national and regional variations in the burden were seen for all these diseases. Public health policy- and decision-makers should devise and implement more effective and geo-specific strategies aimed at counteracting and mitigating the future burden of these diseases. Further research is also warranted to expand our knowledge of potential risk factors associated with MC, AC and OC and to improve the prevention, early detection and treatment of these diseases.

## Data Availability

All data used in this research are publicly available

http://ghdx.healthdata.org/gbd-results-tool

## Acknowledgments

The authors would like to thank all members of the Institute for Health Metrics and Evaluation (IHME), and all collaborators involved in GBD 2017 study.

## Contributors

HD: involved in data collection, study design, statistical analysis, manuscript preparation and supervision. DL and AAM: involved in study design, data interpretation and manuscript review. AY: involved in data interpretation and manuscript review. YL and NLB: involved in study design, statistical analysis, manuscript preparation, manuscript review and supervision. JW: involved in manuscript review and supervision. All authors have read and approved the final manuscript.

## Patient and public involvement

Patients or the public were not involved in the design, data collection, analyses, or interpretation of this research.

